# A review of socioeconomic inequalities in HIV infections among women in Low and Middle-income countries

**DOI:** 10.1101/2022.07.01.22277077

**Authors:** Adeoluwa P Adegbosin, Adeyinka E Adegbosin

**Affiliations:** Georgia State Department of Community Health, Atlanta, Georgia; School of Medicine, Griffith University, Gold Coast, Queensland

## Abstract

**Background:** Evidence shows that HIV is more prevalent among women in low and middle-income countries, this disparity is partly due to biological and socioeconomic factors. In this study we synthesised evidence on the relationship between socioeconomic factors and HIV prevalence among women.

**Method:** We searched PubMed, Cochrane, Scopus, and Embase, for articles published between 2010 and 2020 that investigated the relationship between socioeconomic factors and HIV prevalence among women in low and middle-income countries. We analysed and reported our findings based on the PRISMA guidelines.

**Findings:** A total of 14 studies were included in the final analysis. Six studies explored the effect of disparity in level of education on the prevalence of HIV among women. The effect of income disparity was explored in eight studies, while two studies each explored the effects of occupation and employment status. Overall, evidence shows that women with low socioeconomic status were more at risk of HIV infection.

**Interpretation:** Targeted Interventions aimed at improving the socioeconomic status of women in low and middle-income countries will play a significant role in reducing the prevalence of HIV infection in these settings.

## Introduction

A review of the existing studies shows that the number of women living with HIV is relatively higher than that of men living with HIV. (1,2,3) According to Rodrigo and Rajapakse (2), the trend changed in the late 2000s when the percentage of women with HIV reached 50 percent of the people living with HIV, compared to 1985 when it was only 35 percent. Since then, the trend has been on an upward trend. In 2011, the UNAIDS estimated that it was about 58 percent of the adults living with HIV (3). In 2016, approximately 17.8 million people out of the 34.5 million people, about 52 percent of the people living with HIV globally, were females aged above 15 years. Although evidence varies from one study or year to the other, it suggests that the percentage of women living with HIV is relatively higher than that of the male counterparts. According to Abbai, Wand, and Ramjee (1), it is about 60 percent in Africa, suggesting that it has been increasing rather than decreasing.

The higher HIV prevalence among women is attributed to many factors. According to Hegdahl, Fylkesnes, and Sandoy (4), women are more susceptible to HIV due to their biological factors that expose them to the disease during sexual intercourse. According to Girum et al. (5), unequal power relations, violence against women, and sexual coercion are among the major factors contributing to high HIV prevalence among women than among men. The unequal power relations give men excessive power over women and may result in sexual coercion and violence against women, increasing their exposure to HIV-related infections. According to Naicker et al. (6), sexual behaviors such as multiple sex partners are also contributing to the high HIV prevalence among women.

Adding to the higher HIV prevalence among women than men is the socioeconomic inequalities between the two groups. According to Hegdahl, Fylkesnes, and Sandoy (4), age discrepancies in relationships, limited access to education, and gender-based violence are among the socio-demographic factors that expose women to higher HIV prevalence than men. Socioeconomic differences between the urban and rural dwellers have also been attributed to higher HIV prevalence among women. Poverty has also been identified as one of the major factors contributing to high HIV prevalence among women. According to Igulot and Magadi (7), it exposes women to HIV infection by denying them access to education, loss of livelihood, and early marriages, among other things.

Some studies have also found a bi-directional correlation between HIV prevalence and wealth/poverty. According to Bunyasi and Coetzee (8), poverty has both a downstream and an upstream effect on HIV prevalence. With regard to an upstream effect, the low-income families, including women who are identified as relatively poorer than men, are likely to engage in age-disparate or transactional sexual relationships that expose them to higher chances of HIV infection. Regarding the downstream effect, the HIV infection is likely to impoverish the people who succumb to the disease and also worsen their economic conditions.

In addition, Hajizadeh et al. (9) showed that poverty may constrain the ability of poorer women, to negotiate for safe sex practices, including the use of a condom. There is no existing systematic review on the effect of inequality on HIV prevalence among women, hence this review is a synthesis of evidence from existing studies on socioeconomic inequalities and HIV prevalence among women living in low and middle-income countries.

The review explores socioeconomic inequalities with regards to occupational backgrounds, social class, income, neighbourhood deprivation, and educational achievements (10).

## Methods

### Search strategy

The systematic review was conducted between April 2021 and May 2021 to identify the articles included in the analysis. Four databases: PubMed, Cochrane, Scopus, and Embase were searched using the following key search terms: Inequality OR Inequity OR/AND Equity AND HIV AND Low-income countries OR/AND Middle-income countries OR/AND Developing countries. The search date was limited to between 2010 and 2020 to ensure that only the latest articles were included in the analysis.

### Selection criteria

Included articles focused on socioeconomic inequality among women from either low-income or middle-income countries, published between 2010 and 2020, and published in a peer-reviewed journal. Additionally, it had to be published in English and a full text. The articles were also either to be systematic review, case-control, narrative, cohort or cross-sectional.

The articles published in other languages, those evaluating HIV on other groups of people other than women and those published in 2009 and beyond were excluded from the analysis. Additionally, those published in developed countries and studies with designs other than a systematic review, case-control, narrative, cohort, and cross-sectional were excluded from the analysis.

### Quality assessment

A checklist developed by the Critical Appraisal Skills Programme (CASP) was used to evaluate the quality of the articles included in the analysis. The checklist consisted of 8 major questions designed to evaluate the validity and strength of the articles. Every article was graded based on the extent to which it fulfilled or did not fulfil the checklist’s criteria. A (++) symbol was used to indicate that the article fulfilled all or most of the criteria, (+) was used to indicate that it fulfilled some of the criteria, whereas (-) was used to indicate that it met a few of the criteria. Table 1 summarizes the outcomes of the quality assessment for the articles included in the review.

**Table 1:**
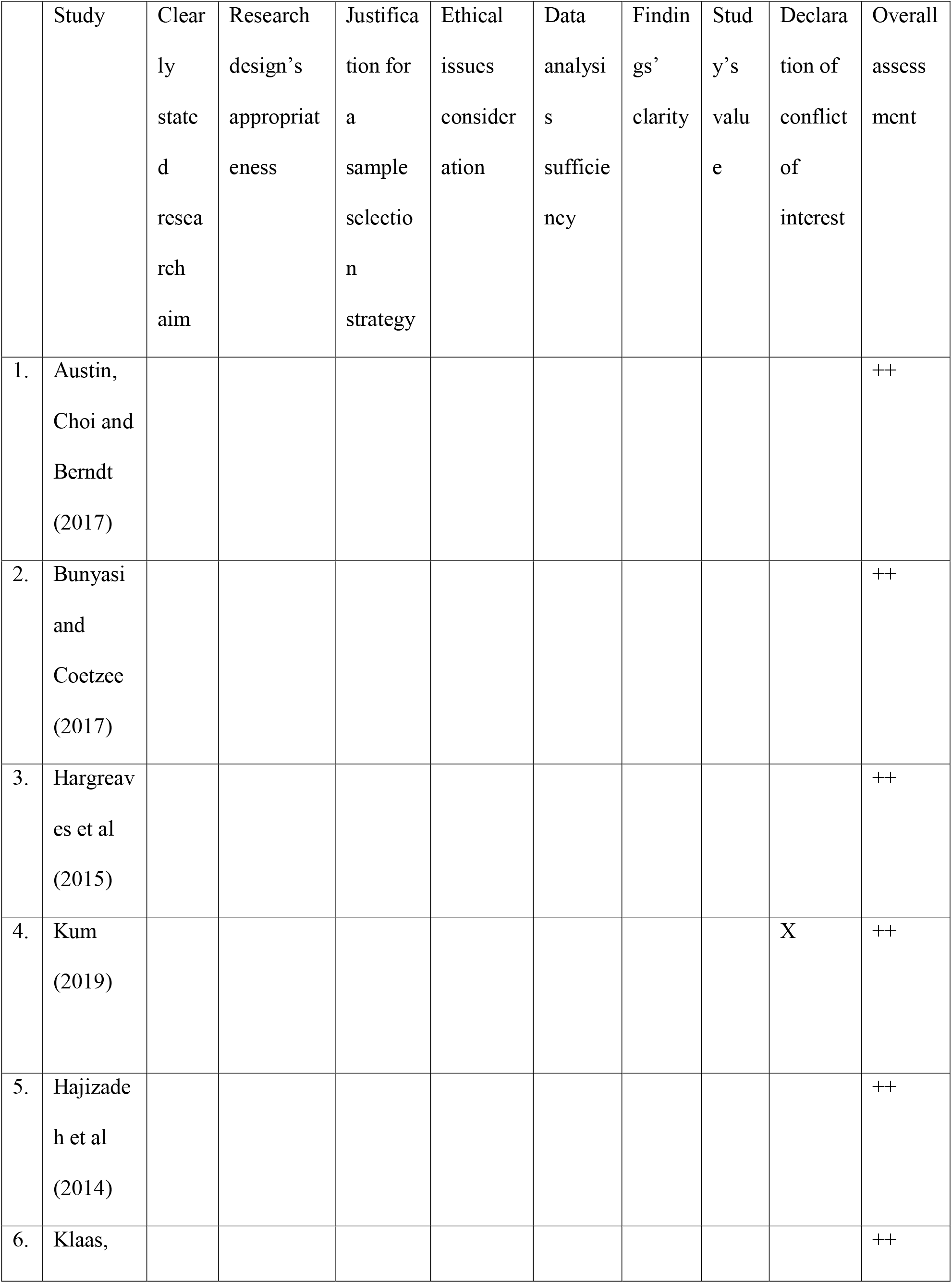
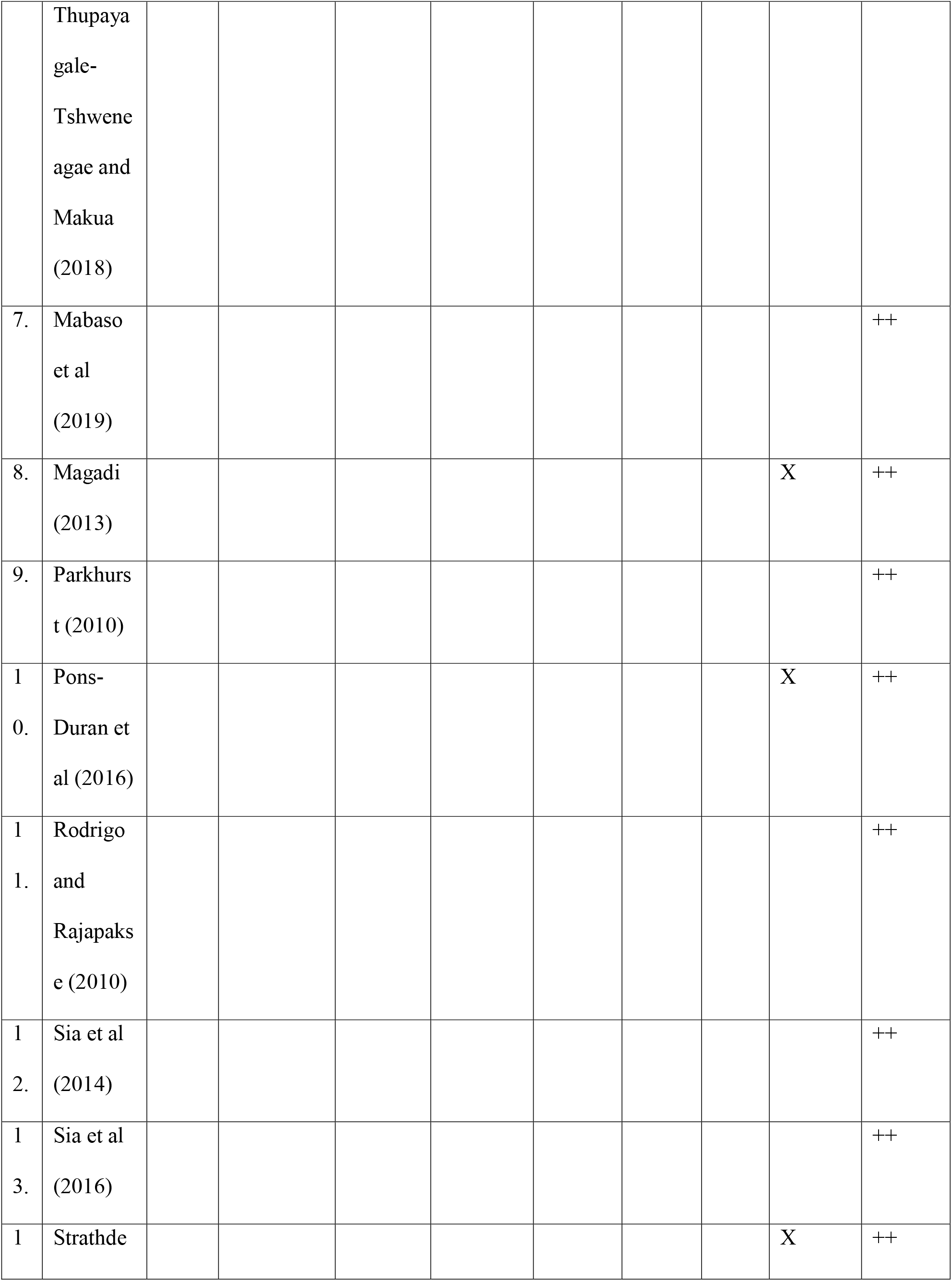
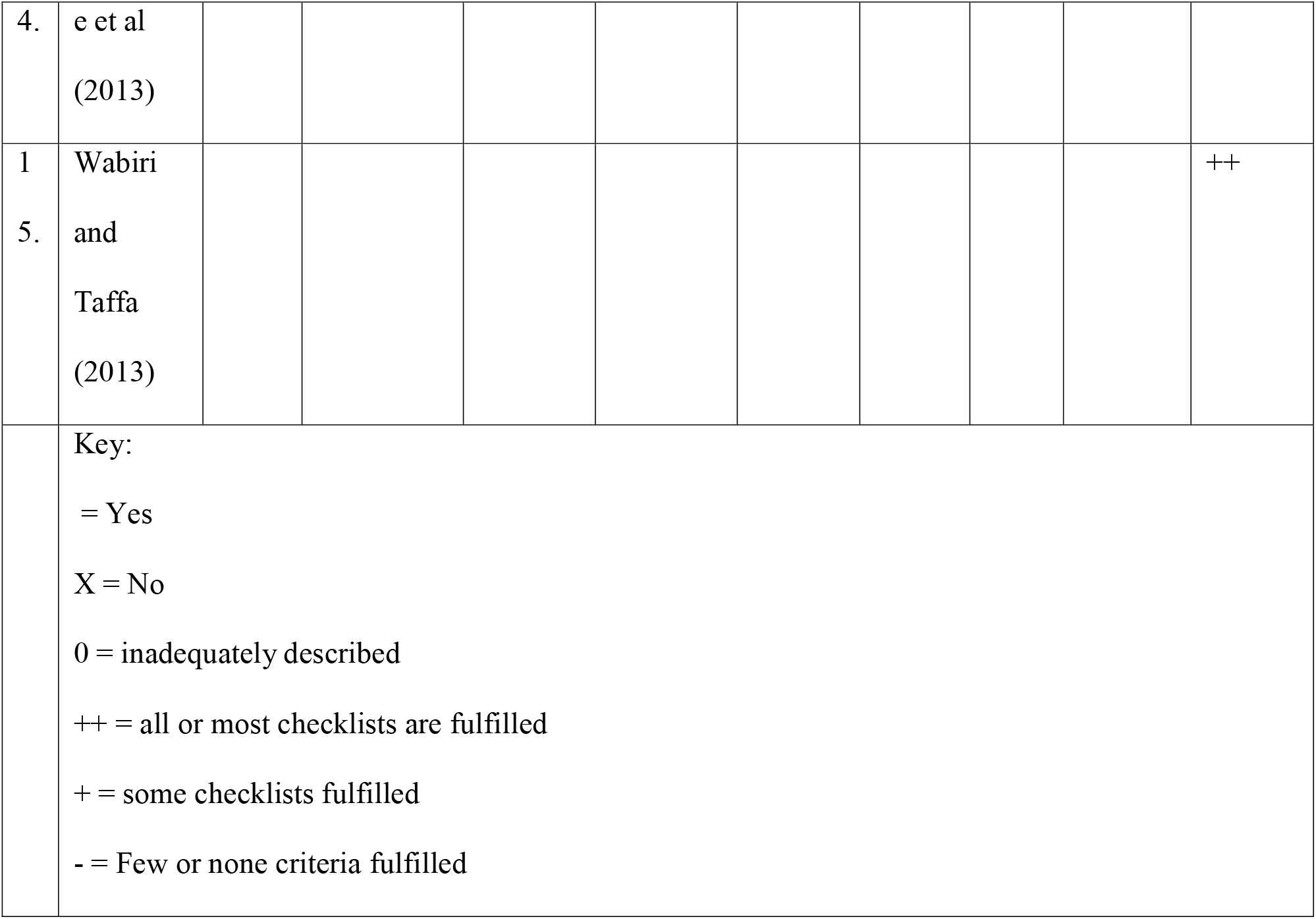
Quality assessment of included articles.

## Results

### Study selection process

A total of (519) articles were identified through database searching whereas (79) more articles were identified from other sources (Figure 1). The articles were identified based on their titles and areas of focus, which was socioeconomic inequality among women. 231 duplicate articles were removed from the analysis. A total of (367) articles were removed from the review because they were unrelated to socioeconomic inequalities among women living in low and medium-income countries. This left total of (231) articles for further evaluation. The full-text evaluation excluded (216), a total of (14) articles met the inclusion criteria. Table 1 provides the quality assessment for the articles that met the inclusion criteria.

**Figure 1:** The PRISMA diagram depicting the process followed to select the articles

## Discussion

Significant level of socioeconomic inequality was found among women living in low and middle-income countries. Some of this inequality may be attributable to socio-cultural factors in many of the countries included in the review, the inequalities were found to be linked to higher HIV prevalence among women. Nonetheless, the findings differ from one country to the other and from one variable to the other. This part of the review critically synthesizes findings from previous studies to describe the extent to which the inequalities contribute or do not contribute to higher HIV prevalence among women living in low- and middle-income countries. The review is organised based on the different dimensions of inequalities identified from the previous studies.

### Education

This part of the review evaluates the extent to which the differences in the number of years in school or years of formal education or lack of it contribute or do not contribute to HIV prevalence among women included in the review. The general expectation is that HIV prevalence among various groups of people, including women, decreases with increased formal education years. However, the reviewed articles provide mixed results showing that education may not necessarily decrease HIV prevalence among women living in low- and middle-income countries.

Bunyasi and Coetzee (8) appraised the link between HIV infection and socioeconomic status in South Africa. Using the logistic regression analysis and a sample of 1,906 women at reproductive age, they established that every additional year of formal education was linked to a 10 percent decrease in the transmission of HIV from mothers to children in the Western Cape Provinces (WCP), but no such association was established in the Free State Provinces (FSP). Contrary to the studies that link higher levels of education with decreased HIV infection among women, Bunyasi and Coetzee (8) showed that although higher levels of education decreased the rate of infection in WCP, it did not decrease it in FSP. The findings suggest that other factors other than education inequality contributed to HIV reduction in FSP.

Mabaso et al. (11) evaluated the gender and other related factors that contributed to the difference in HIV prevalence among different groups of people in South Africa. Using a sample of 86,000 participants, they established that tertiary education was among the factors that reduced HIV prevalence among black South African women. This was in congruence with part of Bunyasi and Coetzee findings (8).

Sia et al. (12) evaluated the influence of education, among other factors on HIV prevalence in Tanzania, Lesotho, and Kenya. Using a sample of 51,059 from the three countries, they established that lack of education among women contributed to higher HIV prevalence in Kenya. Similarly, they identified education attainment as a protective factor to women living in Lesotho, implying that education may reduce HIV prevalence among women living in low- and middle-income countries, whereas lack of it may expose them to HIV infection.

Hargreaves et al. (13) evaluated the link between HIV prevalence and socioeconomic inequalities among young people living in seven African countries. Using a sample of between 2,408 and 12,082 participants from each country, they established that HIV prevalence was relatively higher among the educated women in Malawi and Ethiopia, but it was relatively lower among their counterparts from Zimbabwe, Kenya, and Lesotho. On the one hand, the study’s findings were in agreement with Bunyasi and Coetzee (8), Mabaso et al. (11), and Sia et al. (12) who identified education attainment as a way to decrease HIV prevalence among women living in low- and middle-income countries. On the other hand, they were in disagreement with them because they identified Malawian and Ethiopian educated women as among the people with higher levels of HIV infection.

The review of the literature shows that in general, education contributes to reducing HIV prevalence among women living in low- and middle-income countries in different ways. According to Kum (14), it provides women with the knowledge and information they require to protect themselves from HIV infection. The information and knowledge in turn, transform their behavioural practices and reduce their chances of engaging in prostitution (15). In view of this, several studies have advocated for educational programs to educate girls to reduce HIV infection among them in the future. While the programs may be long-term, they reduce education inequalities that have been identified as exposing women living in low and middle-income countries to higher HIV prevalence.

Evidence from Malawi and Ethiopia depicts that higher education levels may expose women to HIV infection instead of protecting them. The review of the literature shows that poverty may be a contributing factor to such practices. Young girls from poor families may engage in prostitution to enable them to acquire higher levels of education. While evidence from different authors provides conflicting findings of the influence of education on reducing HIV infection among women, it is possible that poverty forces young women during their processes of acquiring higher levels of education to prostitution or pre-marital sex practices. For this reason, higher education levels may not be directly related to failure to reduce Table 2 summarises the above findings.

**Table 2:** Studies evaluating the effect of education inequality on HIV prevalence among women

| Article | Sample size | Country | Study type | Key finding |
| --- | --- | --- | --- | --- |
| Bunyasi and Coetzee (2017) | 1,906 | South Africa | Cross-sectional | • Every additional year of formal education was linked to a 10 percent decrease in the transmission of HIV from mothers to<br>children in the Western Cape Provinces<br>• No such association was established in the Free State Provinces (FSP)<br>• <u>Implications</u>: other factors other than the level of education reduced HIV prevalence in FSP |
| Mabaso et al. (2019) | 86,000 | South Africa | Cross-sectional | • Identified tertiary education as one of the factors that reduced HIV prevalence among black South African women |
| Sia et al. (2014) | 51,059 | • Kenya<br>• Lesotho<br>• Tanzania | Cross-sectional | • Lack of education among women contributed to higher HIV prevalence among them in Kenya.<br>• In contrast, education attainment was<br>identified as a protective factor to women living in Lesotho<br>• <u>Implication</u>: Education may be effective at reducing HIV prevalence whereas lack of it may expose them to HIV infection. |
| Hargreaves et al. (2015) | Between 2,408 and 12,082 | • Malawi<br>• Ethiopia<br>• Kenya<br>• Lesotho<br>• Zimbabwe | Cross-sectional | • HIV prevalence was relatively higher among the educated women in Malawi and Ethiopia<br>• However, it was relatively lower among their counterparts from Zimbabwe, Kenya, and Lesotho<br>• <u>Implications</u>: Higher levels of education may either expose or prevent women living in low- and middle-income<br>countries from HIV infection |
| Kum (2019) | Unspecified | • Malawi | Cohort | • Education provides women with information and knowledge they require to protect themselves from HIV infection |
| Strathdee et al. (2013) | 3 cases | • Africa<br>• Dominican republic<br>• Mexico | Narrative | • Information and knowledge from education contribute to behavioural changes among women; thus, reduce their probabilities of engaging in prostitution |

### Wealth/income

This part of the review evaluates the extent to which wealth/income levels contribute or do not contribute to higher HIV prevalence among women living in low- and middle-income countries. Bunyasi and Coetzee (8) evaluated the extent to which wealth contributed to HIV prevalence among South African women living in Free State Provinces (FSP) and Western Cape Provinces (WCP). They identified women in the second-lowest wealth quintile from the Western Cape Provinces as highly susceptible to HIV infection. Nonetheless, their counterparts from both provinces at the highest wealth quintile had the least HIV prevalence suggesting that wealth inequality was a major contributor to HIV infection. The study suggests that higher levels of income and wealth may decrease the rate of HIV infection among women.

Wabiri and Taffa (16) evaluated the link between HIV prevalence and socioeconomic inequalities in South Africa. Using a sample of 14,384, they established that the HIV prevalence was highest among women from low-income households and least among their counterparts from a high-income household. The study further established that income levels had a higher influence on HIV testing and access to HIV-related information among the women included in the analysis. Those from low-income households experienced higher stigma than their counterparts from high-income households. In a similar study, Mabaso et al. (11) identified higher socioeconomic status as effectively reducing HIV prevalence among black South African women. Both studies suggest that higher income levels may effectively reduce HIV infection among women living in low and middle-income countries.

Magadi (17) evaluated how poverty contributed to HIV infection among poor people living in urban areas in Africa. The study established that the rate of HIV infection was amplified by poverty among the women living in urban areas. Using health surveys conducted in 20 different African countries. The study suggested that low-income levels among African women living in urban areas could be a health risk among them, thereby exposing them to higher HIV infection.

Studies conducted elsewhere link higher income levels to higher HIV infections among women living in low- and middle-income countries. Hajizadeh et al. (9) conducted a study to evaluate the influence of socioeconomic inequalities on HIV prevalence among African countries linked higher socioeconomic status among women to higher HIV prevalence. Among the 24 African countries included in the analysis, only Senegal and Swaziland had relatively higher rates of HIV infection among women living in poor households. The study suggests that while higher income levels among women may deter higher HIV infection, it may as well be a health risk to them.

Parkhurst (18) evaluated how poverty and wealth contributed to HIV infection among 12 African countries. Using trend analysis and retrospective ecological comparison methods, the study did not establish a consistent trend between household wealth quintile and HIV prevalence among the African countries included in the analysis. It established that HIV prevalence among women increased with wealth increase in Uganda and Ivory Coast. Nonetheless, it decreased among Tanzanian women from wealthy households, but increased among their counterparts from poor families. While the study provides conflicting results suggesting that wealth may either increase or decrease HIV infection among women, it is in agreement with Hargreaves et al. (13) who identify that wealth can influence HIV infection either way. This suggests that different factors, including wealth, may influence HIV infection among women either way. For this reason, effective measures should be developed for individual countries and specific regions as opposed to generalising the measures among women living in low- and middle-income countries.

Pons-Duran et al. (19) appraised the link between socioeconomic status and HIV infection in Mozambique. Using a sample of 1,511 adults, they established that while HIV prevalence was relatively higher among women than among men, the prevalence reduced with an increase in income levels suggesting that the poor women were at higher risks of HIV than the rich women.

In a systematic review of studies evaluating the link between HIV prevalence and poverty among women living in developing countries, Rodrigo and Rajapakse (2) identified poverty as a risk factor that exposed women to HIV infection. They established that the high poverty levels among women denied them access to education, employment opportunities, and health care services. Although each of these factors was not linked directly to HIV infection, they contributed in one way or the other to HIV infection. The lack of education forced women to early marriages and the knowledge they required to protect themselves from HIV infection. The lack of education also decreases their chances of securing employment, which forces them to depend largely on men. The over-dependence on men denies women power over sexual-related issues, thereby exposing them to HIV infection. Table 3 summarises the above findings.

**Table 3:**
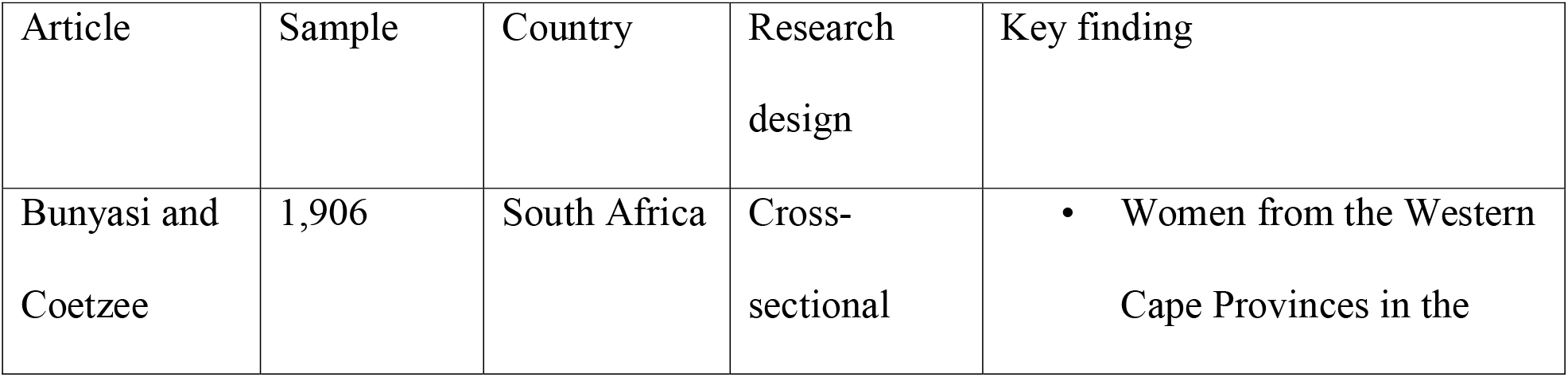
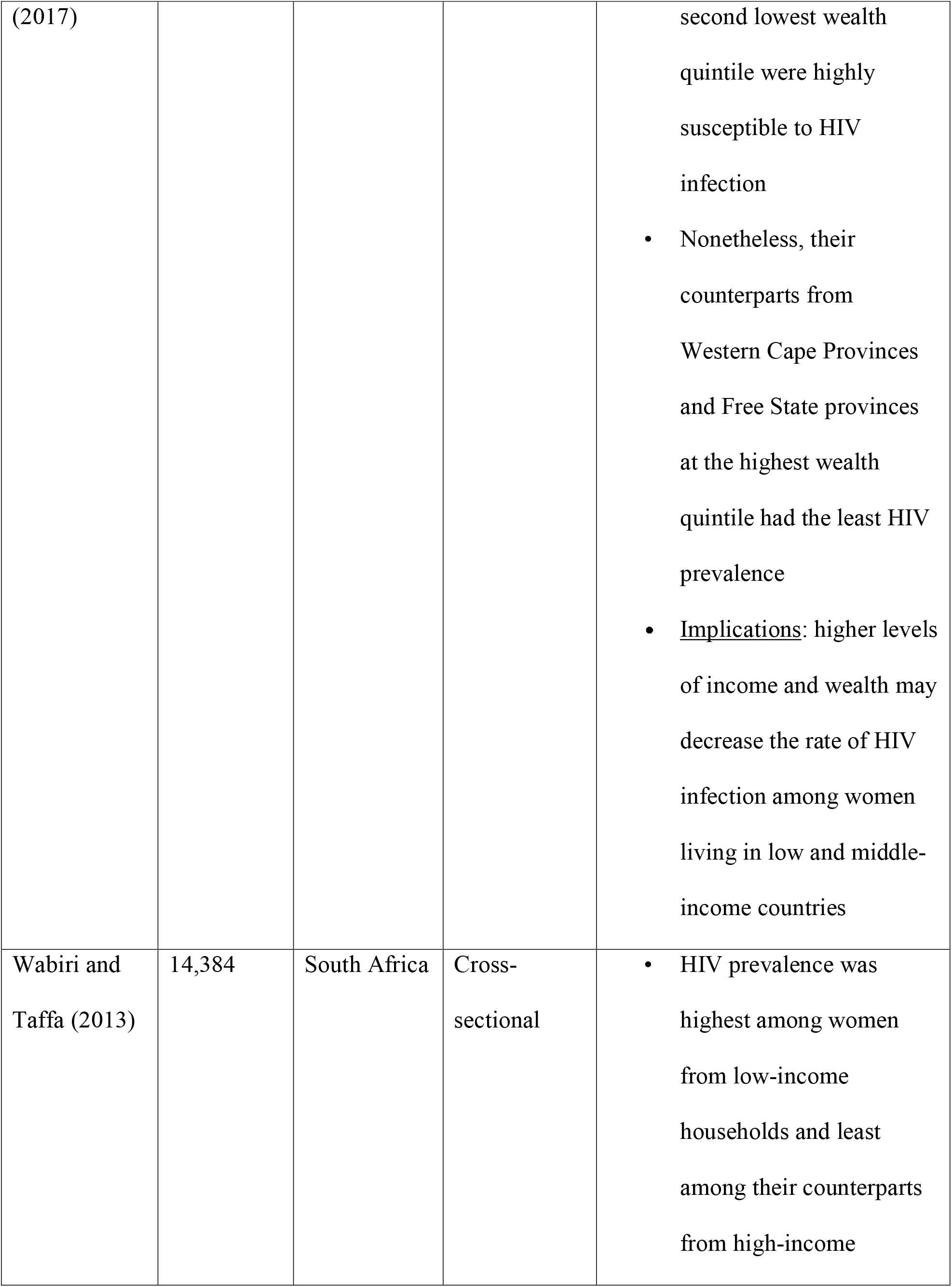
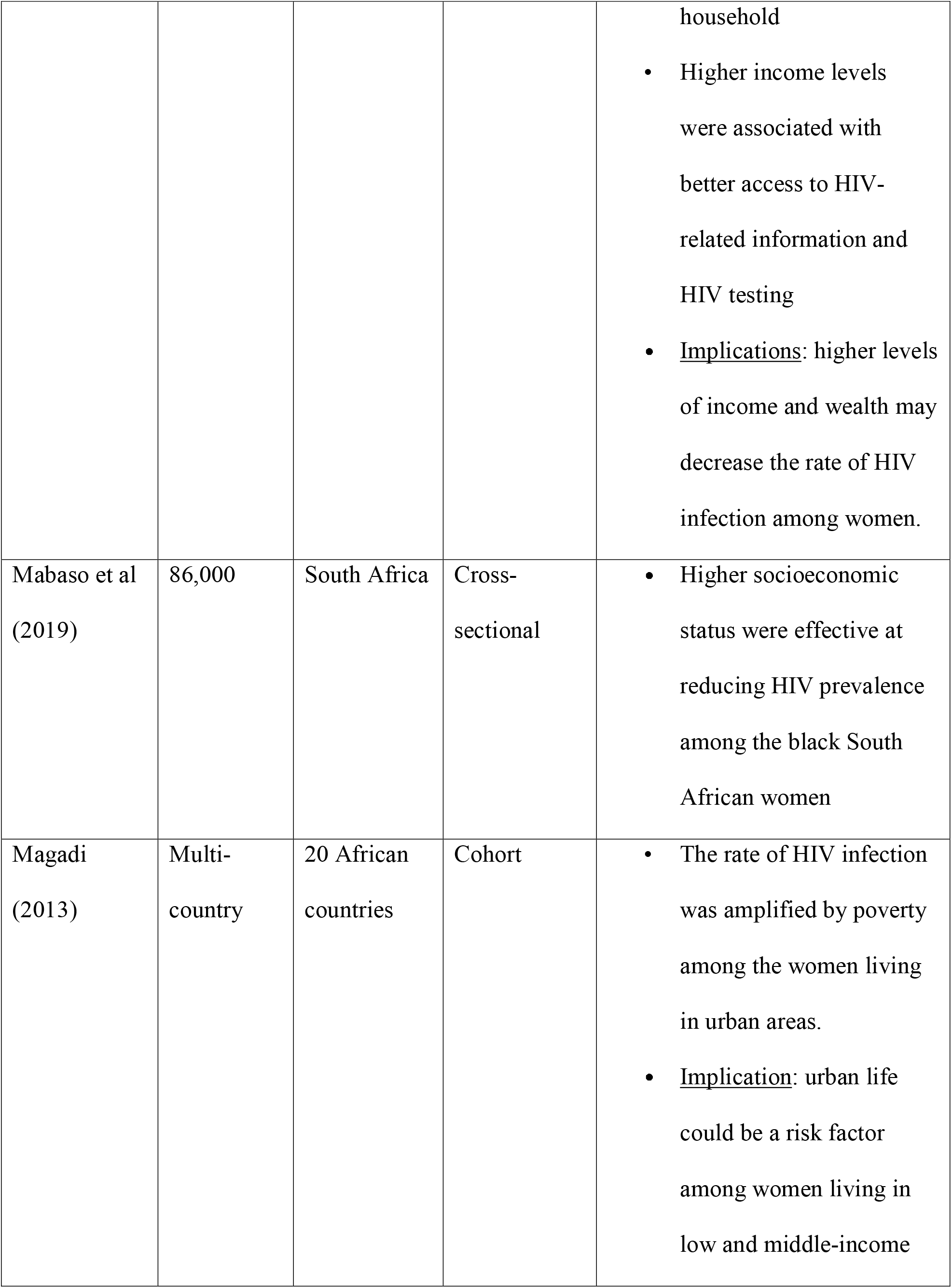
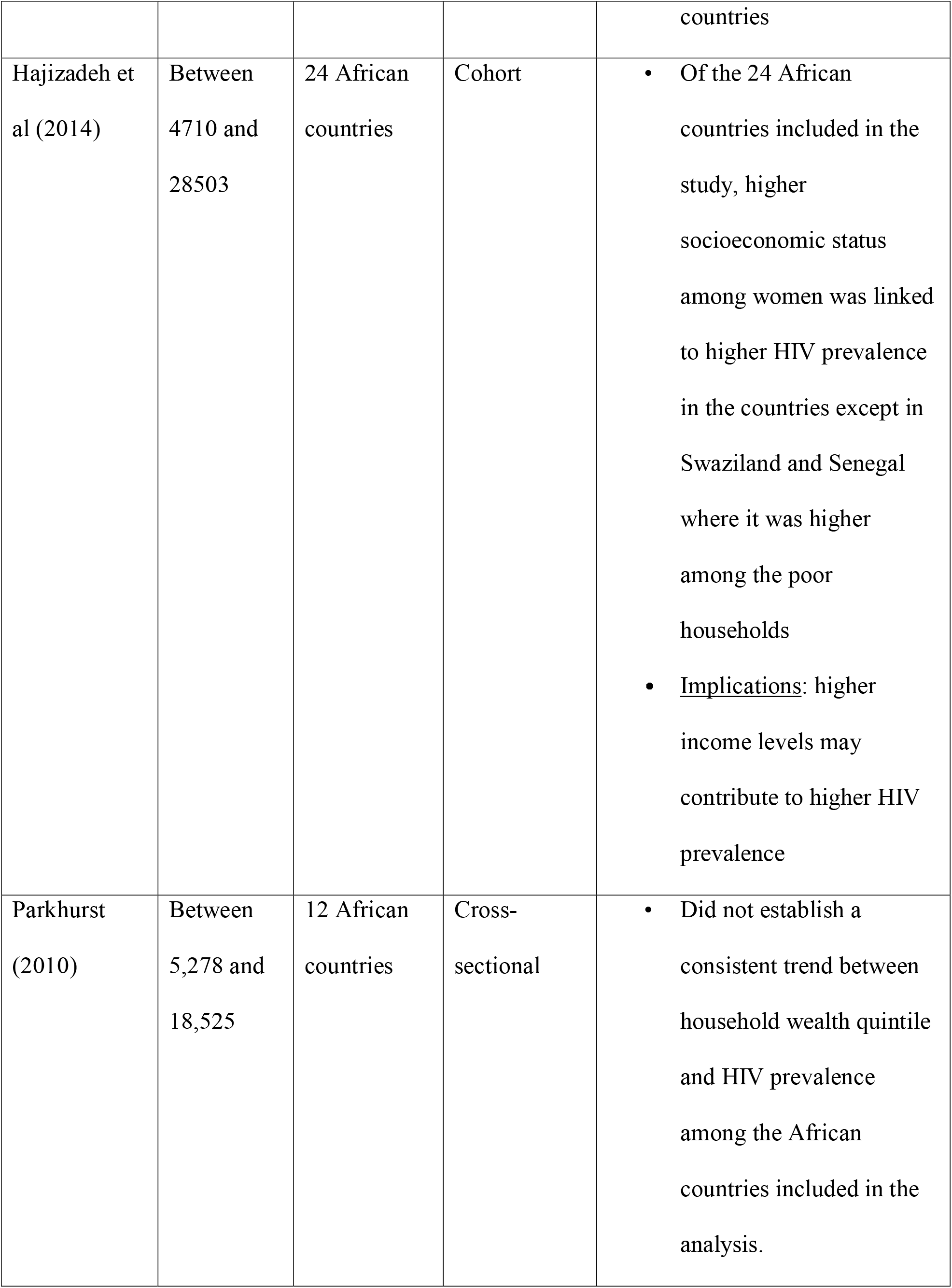
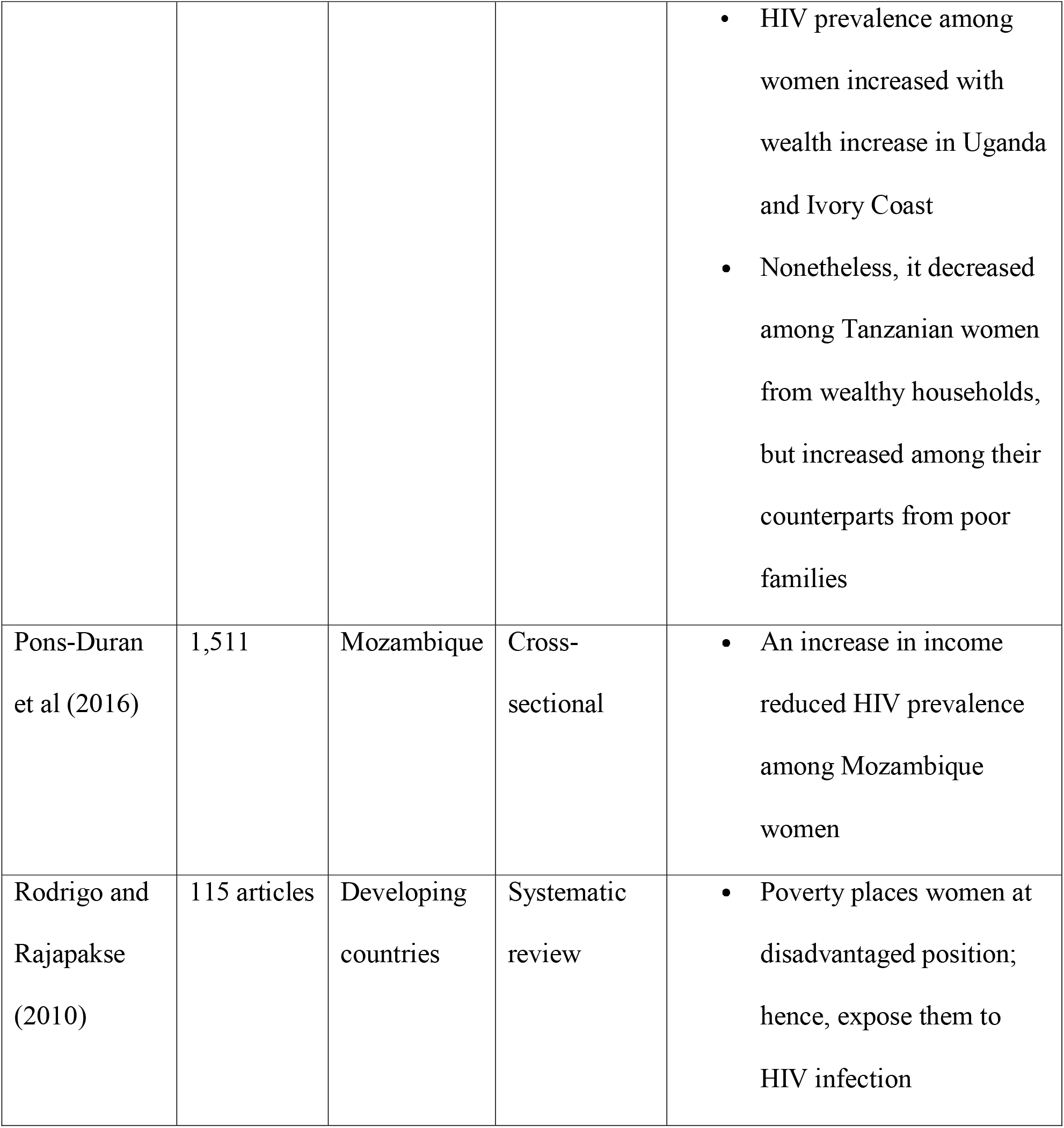
Studies evaluating the effect of wealth/income inequality on HIV prevalence among women

There are several ways that wealth and income contribute to higher HIV prevalence among women living in low- and middle-income countries. According to Rodrigo and Rajapakse (2), poor women living in low- and middle-income countries may be forced into sex by their partners or young women forced into early marriages by parents due to poverty. Similarly, poverty may prevent young female children from going to school; hence, deny them the knowledge to advocate for their rights or know what they ought to do to prevent them from HIV infection. Additionally, poverty may push women living in low- and middle-income countries into unsafe sexual behaviours, which may heighten HIV prevalence among them. According to Strathdee et al. (15), it may force them into prostitution to earn income. Furthermore, it may deny women the power to negotiate for safe sexual practices. (20)

Ramjee and Daniels (21) find that low-income levels may drive young women into early sex experiences, contribute to multiple sex partners, non-consensual sex experiences, and lower condom use, which increases HIV prevalence among women who engage in such practices. Sawers and Stillwaggon (20) found that poverty may force women into transactional sex that exposes them to HIV infection. Poverty may also expose women to different forms of violence, which have been linked to high levels of HIV infections among women than among men. According to Rodrigo and Rajapakse (2), the large-scale civil violence witnessed among some of the developing countries such as Nepal and Ethiopia contribute to the displacement of people, loss of jobs and spouses, which largely affect women, thereby exposing them to high HIV infection. At the family level, the low socioeconomic status may hinder women from leaving violent husbands. Additionally, the high levels of poverty among women may interact with other gender-related inequalities to expose women living in low- and middle-income countries to HIV infection.

There are several ways that wealth and high-income levels reduce HIV prevalence among women living in low- and middle-income countries. According to Wabiri and Taffa (16), it allows women to access HIV-related information and, to some extent, encourages them to go for HIV testing contrary to their counterparts from low-income levels. The information they acquire, in turn, helps them to make informed decisions; thus, decrease HIV infection among women from wealthy households. (14) According to Strathdee et al. (15), it decreases the tendency of women to engage in prostitution and other unsafe sex behaviours that expose them to HIV infection.

### Occupational background

This part of the review evaluates the extent to which the differences in occupational background between men and women living in low- and middle-income countries contribute or does not contribute to HIV prevalence among women included in the review.

Sia et al. (22) evaluated the gender-related factors contributing to differences in HIV prevalence among 21 different African countries. Using health surveys from those countries, they established that the higher HIV prevalence among Rwandese, Mozambican, Ethiopian, and Ivory Coast women than their male counterparts resulted from differences in occupations. The study established that the agricultural related occupations that women worked in exposed them to HIV infection. While such may be a case in isolation, it suggests that the menial jobs that women rely on for income forces them to depend on men, and in so doing, exposed to HIV infection. This may be explained by the fewer alternatives that most of the women living in low and middle-income countries have in comparison to their male counterparts.

Klaas, Thupayagale-Tshweneagae Makua (24) appraised the role that gender played in the spread of HIV among South African farm workers. Using a sample of 22 research participants, the study established that the nature of the farm occupations that the respondents performed exposed women to HIV infection due to lack of power in decision-making processes of issues related to sexual matters. The findings were in agreement with Sia et al. (22), suggesting that the agricultural jobs that women living in low and middle-income countries perform may be a contributing factor to HIV infection. Table 4 summarises the findings.

**Table 4:** Studies evaluating the effect of occupational inequality on HIV prevalence among women

| Article | Sample | Country | Research design | Key finding |
| --- | --- | --- | --- | --- |
| Sia et al (2016) | Between 2,000 and 10,000 | 21 African countries | Cohort | • Higher HIV prevalence among Rwandese, Mozambican, Ethiopian and Ivory Coast women than their male counterparts resulted from differences in occupations.<br>• The study established that the agricultural-related occupations that women worked in exposed them to HIV infection.<br>• Implication: the nature of the jobs that women perform contribute to<br>higher HIV prevalence among them |
| Klaas, Thupayagale-Tshweneagae and Makua (2018) | 22 | South Africa | Narrative | The nature of farm occupations that women performed denied them power over sexual related issues; hence, exposed them to HIV infection |

The above results suggest that most of the women living in low- and middle-income countries lack power in most of the sexually related issues due to the nature of the work they perform. In addition, they indicate that due to the few occupation alternatives, most of the women rely on men for most of their basic needs; as such, they may be more exposed to HIV infection.

### Employment status

Austin, Choi, and Berndt (24) appraised the influence of high unemployment among young women aged between 15 and 24 years living in developing countries. The study established that lack of formal employment contributed to transactional sex, thereby exposing them to HIV infection. The results suggested that the nature of employment among women was a major contributor to HIV infection among unemployed women. However, the study did not identify employment among the young women as reducing HIV infection among them. Table 5 summarises the findings.

**Table 5:** Studies evaluating the effect of employment status inequality on HIV prevalence among women

| Article | Sample | Country | Research design | Key finding |
| --- | --- | --- | --- | --- |
| Bunyasi and Coetzee (2017) | 1,906 | South Africa | Cross-sectional | • Employment status is not linked to HIV prevalence among women in both Free State provinces and Western Cape provinces |
| Austin, Choi and Berndt (2017) | Unspecified | African countries | Cross-sectional | • Unemployment exposed young women aged between 15 and 25 years to HIV infection<br>• Employment did not reduce HIV infection suggesting that other factors such as transactional sex and concurrent partners contributed to HIV infection among<br>• Implication: Other mechanisms other than<br>employment should be<br>used to reduce HIV<br>infection among young<br>women |

Overall, this review showed that in addition to biological factors, socio-disparities in income, education, and occupation amongst other factors, contribute to a higher prevalence of HIV among women living in low and middle-income countries.

## Data Availability

All data produced are available online on PubMed, Cochrane, Scopus, and Embase.

