## Supplementary material for "A review of socioeconomic inequalities in HIV infections among women in Low and Middle-income countries": Figure 1

**Screening**

**Included**

**Eligibility**

**Identification**

Records identified through database searching
 (n = 64)

Additional records identified through other sources
(n = 0 )

Records after duplicates removed
(n = 59)

Records screened
(n = 60)

Records excluded with reasons
(n = 46)

Outside MNCH scope (n = 33)

Conference abstract, No full text (n = 1)

Review articles (n = 2)

Study setting is a developed country (n = 7)

Not peer reviewed (n = 2)

Study protocol (1)

Full-text articles assessed for eligibility
(n = 14 )

Full-text articles excluded, with reasons
(n = 0 )

Studies included in qualitative synthesis
(n = 18 )
